# Analysis of serum trace elements, macro-minerals, antioxidants, malondialdehyde and immunoglobulins in seborrheic dermatitis patients: A case-control investigation

**DOI:** 10.1101/2020.10.01.20205534

**Authors:** Ishrat Jahan, Md. Rabiul Islam, Md. Reazul Islam, Rubaiya Ali, S. M. Matiur Rahman, Zabun Nahar, Abul Hasnat, Md. Saiful Islam

## Abstract

**Background:** There are many theories abound; the exact mechanism for the pathophysiology of seborrheic dermatitis (SD) remains unknown. Neuropsychiatric disorders, stress, weakened or irregular immune responses, fungal infections, etc. are thought to be associated with the development of SD. The present study aimed to determine the serum levels of trace elements (TEs), macro-minerals (MMs), antioxidant vitamins, malondialdehyde (MDA), and immunoglobulins in SD patients to explore their role in the disease progression.

**Methods:** This prospective case-control study recruited 75 SD patients and 76 healthy controls (HCs) matched by age and gender. Serum TEs and MMs were analyzed by the flame atomic absorption spectroscopy and graphite furnace atomic absorption spectrometry. RP-HPLC was used to determine the serum levels of vitamin A and E. Serum MDA levels were measured by UV spectrophotometry as a marker of lipid peroxidation, whereas the turbidimetric method was used to quantify the serum levels of immunoglobulins.

**Results:** We found significantly higher concentrations of serum copper, manganese, and iron, calcium, and magnesium in SD patients compared to HCs. Serum levels of vitamin E significantly decreased and serum levels of MDA significantly increased in SD patients. Besides, the lower concentrations of immunoglobulin A, G, and M were observed in SD patients when compared to HCs. The present study also found a positive correlation between serum Zn and Ca levels (*r* = 0.365, *p* = 0.009) in SD patients; whereas a negative relation was detected between serum Cu and Ca levels (*r* = -0.298, *p* = 0.035).

**Conclusions:** The present study suggests that increased levels of TEs, MMs, MDA, and decreased levels of immunoglobulins and vitamin E are strongly associated with the pathophysiology of SD. Moreover, these alterations may provide a predictive tool for the assessment and management of SD patients.

## Background

Seborrheic dermatitis (SD) is one of the most common skin disorders and a large number of people suffer from this skin disease [1]. This chronic condition also termed as papulosquamous dermatosis since both scales and papules have been found notably. The affected skin becomes edematous, pinkish, and covered with yellow-brown scales and crusts [3]. Both infants and adults can be affected by SD [2]. An excess amount of sebum is produced from the scalp; sebaceous follicle-rich areas on the face and trunk are seen in SD patients. It is thought to be one of the most common skin manifestations in patients with human immunodeficiency virus (HIV) infection [4]. There is no data available on the exact incidence of SD in infants, but the disorder is common [5]. The prevalence of SD in adults is believed to be more common than psoriasis. In the USA, the prevalence of SD among the general population is at least 3-5% [6]. Men are affected more often than women in all age groups [7]. Approximately, 15-20% of the total population is suffering from dandruff, which is usual and thought to be the mildest form of SD [8]. In Bangladesh, the prevalence of SD is in a similar magnitude as in developed countries. Statistics show that 15.5 % of populations are suffering from skin diseases in Bangladesh [9]. Several factors such as hormone levels, fungal infections, nutritional deficits, neurogenic factors may be responsible for the development of skin diseases [10]. An inadequate amount of work has been done to reveal the possible reasons for the development of SD. Further knowledge about SD is required for the design of new drug molecules against this skin disease.

Inorganic minerals and nutrients present in all body tissues are vital for the maintenance of normal body functions [11]. The importance of sodium, chloride, and calcium in SD has been well recognized for different biochemical processes and immunological functions [12, 13]. Moreover, different trace element (TE) and macro-mineral (MM) such as copper (Cu), iron (Fe), manganese (Mn), zinc (Zn), calcium (Ca), magnesium (Mg) can induce cellular oxidative stress by various mechanisms [14]. According to the American Academy of Dermatology (AAD), three-dimensional structures of many proteins are important for healthy skin. Some TEs and MMs have a function to maintain this three-dimensional structure of proteins [15]. Elevated levels of these TEs and MMs are considered to be responsible for developing various skin diseases [16]. Altered serum Zn levels were observed in SD patients compared to control subjects [17]. Therefore, changes in serum TEs and MMs levels may influence the pathophysiology and development of SD.

Human skin gets constant exposure to oxidative stress induced by reactive oxygen species (ROS) [18]. Oxidative damages such as DNA modification, secretion of inflammatory cytokines and lipid peroxidation, etc. are usually caused by ROS [19]. Physiological defenses against these have been shown by a series of enzymatic and non-enzymatic antioxidants by inactivating free radical [20, 21]. Natural antioxidants like vitamins A and E are considered to be vital free radical eliminator due to their lipophilic nature since synthetic antioxidants may have some detrimental effects on our body [22,23,24,25]. Conversely, MDA is the end product of lipid peroxidation, which serves as a marker for the oxidative status of the biological system and worth measuring [26].

Immunoglobulins are protein macro-molecules produced by blood cells and neutralize various pathogenic bacteria and viruses [27]. Although, a direct correlation with immunoglobulins and skin diseases is yet to be determined; the relationship between immunodeficiency and SD has been reported by several studies, where SD is found to have distinct clinical and histological features in patients suffering from immunodeficiency virus infection [28-30]. Particularly, the serum Immunoglobulin E (IgE) level is found to be strongly associated with atopic dermatitis [31].

Unfortunately, the knowledge related to the altered serum levels of these parameters in SD is deficient; particularly no such work has yet been done in Bangladesh. Thus, the present study aimed to investigate the serum levels of TEs, MMs, antioxidant vitamins, MDA, and immunoglobulins in SD patients and corresponding healthy controls (HCs) to explore the possible pathogenic cause.

## Methods

### Study population

A total of 151 subjects, 75 SD patients, and 76 HCs were recruited for this case-control study. Patients were recruited from the Department of Dermatology and Venereology, Ad-din Medical College Hospital, Dhaka, Bangladesh. Age and gender-matched (1:1) HCs were taken from different areas of Dhaka city. The subjects who had been treated with any antioxidants that could interfere with the concentrations of trace elements or macro-minerals were excluded from this investigation. Subjects having a previous history of skin disease or pro-inflammatory diseases were also excluded from HCs. Furthermore, the socio-demographic profile of the participants was recorded by using a pre-designed questionnaire. Different biophysical characteristics such as weight, height, and body mass index (BMI) were also analyzed for the study subjects. All procedures were conducted following the principles of the Declaration of Helsinki and later amendments [32].

### Blood collection and preparation

Five-milliliter of the blood sample was collected from the cephalic vein from each participant after an overnight fast. The collected blood samples were then transferred into a metal-free, pre-cleaned test-tube. The samples were permitted to clot for one hour at room temperature (25 °C). Serum samples were extracted from the blood samples by centrifugation at 3000 rpm for 15 minutes and placed into micro-tubes. The separated serum samples were stored at -80 °C until further analysis. Sample collection and preservation was performed according to our previous studies [33].

### Chemicals and reagents

We purchased standard Cu, Fe, Mn, Zn, Ca, and Mg from Buck Scientific, USA. The immunoglobulin kit was purchased from Chronolab, Switzerland. Standard α-tocopherol (vitamin E), α-tocopheryl acetate, and retinol (vitamin A) were supplied by Sigma Chemical Co., USA. For HPLC-based analysis, Active Fine Chemicals Limited, Dhaka, Bangladesh supplied HPLC grade chemicals and reagents. Other supportive chemicals of the recommended grade were purchased from Merck, Germany.

### Determination of serum trace elements and macro-minerals

Flame atomic absorption spectrometry (FAAS) and graphite furnace atomic absorption spectrometry (GFAAS) were used to quantify serum TEs (Zn, Cu, Mn, and Fe) and MMs (Ca and Mg). Briefly, serum samples were diluted with de-ionized water 1:10 dilution. For the calibration curve, different concentrations (0.5, 1.0, 2.0, 5.0 and 10.0 mg/L) of TEs and MMs were used. Finally, the concentrations of TEs and MMs were measured at 422.7, 285.2, 327.4, 248.3, 279.8, and 213.9 nm for Ca, Mg, Cu, Fe, Mn, and Zn, respectively. Standard solutions were run for every 10 test samples to ensure assay accuracy and quality [34]. Enough precaution was taken during the handling of serum samples to avoid or minimize the contamination.

### Measurement of serum antioxidant vitamins

Vitamin A and E were determined from serum samples by liquid-liquid extraction using n-hexane and dried by evaporation using a sample concentrator (DB-3, Techne, UK) at 40°C under a stream of nitrogen. Afterward, the dried extract was reconstituted in the mobile phase. The serum concentrations of vitamin A and E were measured simultaneously at 291nm by the modified RP-HPLC method with UV detection [35].

### Quantification of serum malondialdehyde

The concentration of serum malondialdehyde (MDA) was quantified using thiobarbituric acid (TBA) reagent and the absorbance of the supernatant was measured spectrophotometrically at 530 nm. MDA was measured in terms of TBARS by the modified method described in our past study [33]. The concentration of MDA was expressed as nmol/mL.

### Serum Immunoglobulin profiling

Serum immunoglobulins were estimated by the quantitative turbidimetric method using an immunoglobulin kit. In this method samples containing IgA, IgG, IgM were mixed with the activation buffer and then with anti-human immunoglobulin reagent according to the procedure described in our previous article [36]. Briefly, the serum samples were diluted to 1:4 dilution with saline water. The diluted serum sample was pipetted into a microtitre plate. For calibration curves, separate microtitre plates were used for each of the immunoglobulins (IgA, IgG, IgM). 10 μL, 25 μL, 50 μL, 75 μL and 100 μL of the calibrator protein were also pipetted into marked wells. 230 μL of reagent R1 (tris-buffer) was added to each serum and calibrator protein-containing well of the microtitre plate to make the total volume of 240 μL. Then 15 μL diluted respective anti-human IgA, IgG, and IgM (1:1:1 diluted with saline water) were added to the respective wells and incubated for 2-5 minutes to react with anti-human immunoglobulin with the test serum and calibrator protein. After proper mixing absorbance was taken at 630 nm for IgA and IgG and 405 nm for IgM, respectively.

### Statistical Analysis

All the data were presented as mean ± standard error mean (mean ± SEM). Pearson’s correlation analysis was applied to obtain the correlation among various study parameters. A comparison of all parameters between SD patients and HCs was performed by the independent sample t-tests, Pearson’s correlation test, and box plot graphs. In all cases, a p-value of less than 0.05 was considered as statistically significant. All the statistical analysis was done using the statistical software package SPSS version 25.0 (Armonk, NY: IBM Corp.).

## Results

### Anthropometric and demographic profile of the study population

The anthropometric and demographic characteristics of the SD patients and HCs are presented in Table 1. Where the male compromised the highest percentage for both groups. Though SD initiates at any point of age, our present study was carried out over the patients and HCs with an average age of 18-35 years. There was no significant difference in the mean value of age and BMI between the SD patients and HCs.

**Table 1.** Anthropometric and demographic profile of the study population

| Characteristics | SD patients (n = 75)<br>Mean ± SEM | Healthy controls (n = 76)<br>Mean ± SEM | *p* value |
| --- | --- | --- | --- |
| Age (years) | 27.13 ± 1.61 | 26.30 ± 0.92 | 0.289 |
| BMI (kg/m<sup>2</sup>) | 22.65 ± 1.57 | 22.93 ± 1.31 | 0.432 |
| Sex | Value, n (%) |  |  |
| Male | 43 (57.33%) | 41 (54.67%) |  |
| Female | 32 (42.67%) | 34 (45.33%) |  |

*p* is significant at the 0.05 level.
SD seborrheic dermatitis, SEM standard error mean.

### Serum trace elements, macro-minerals, antioxidant vitamins, MDA and immunoglobulins levels

Serum levels of TEs, MMs, antioxidant vitamins, MDA, and immunoglobulins are shown in Table 2. The concentrations of Cu, Mn, Fe, Ca, and Mg were found significantly increased in the SD patients compared with the HCs. Also, serum MDA levels were found significantly elevated in the SD patients compared to HCs. However, serum levels of vitamin E along with three immunoglobulins (IgA, IgG, and IgM) were observed significantly lower in SD patients compared to HCs. The distribution pattern of different TEs, MMs, antioxidant vitamins, MDA, and immunoglobulins are presented by Fig. 1 and Fig.2.

**Table 2.** Serum levels of trace elements, macro-minerals, antioxidant vitamins, malondialdehyde and immunoglobulins in the study population

| Parameters | SD patients (n = 75)<br>Mean ± SEM | Healthy controls (n = 76)<br>Mean ± SEM | *p* value |
| --- | --- | --- | --- |
| Zinc (mg/L) | 1.430 ± 0.11 | 1.308 ± 0.09 | 0.289 |
| Copper (mg/L) | 2.136 ± 0.10 | 0.950 ± 0.05 | < 0.001* |
| Manganese (mg/L) | 15.830 ± 0.85 | 7.022 ± 0.46 | < 0.001* |
| Iron (mg/L) | 2.248 ± 0.14 | 1.130 ± 0.11 | < 0.001* |
| Calcium (mg/L) | 185.040 ± 5.89 | 99.580 ± 1.77 | 0.001* |
| Magnesium (mg/L) | 29.640 ± 0.55 | 20.640 ± 0.27 | < 0.001* |
| Vitamin A (μmol/L) | 0.403 ± 0.07 | 0.374 ± 0.03 | 0.707 |
| Vitamin E (μmol/L) | 5.564 ± 0.39 | 7.074 ± 0.37 | 0.009* |
| MDA (μmol/L) | 0.2125 ± 0.31 | 0.186 ± 0.28 | 0.011* |
| IgA (mg/dL) | 212.96 ± 16.38 | 263.08 ± 10.22 | 0.001* |
| IgG (mg/dL) | 1208 ± 16.37 | 2317 ± 10.23 | < 0.001* |
| IgM (mg/dL) | 119.45±8.57 | 158.89±9.09 | 0.003* |

\**p* is significant at the 0.05 level.
SD seborrheic dermatitis, SEM standard error mean.

**Fig. 1.**
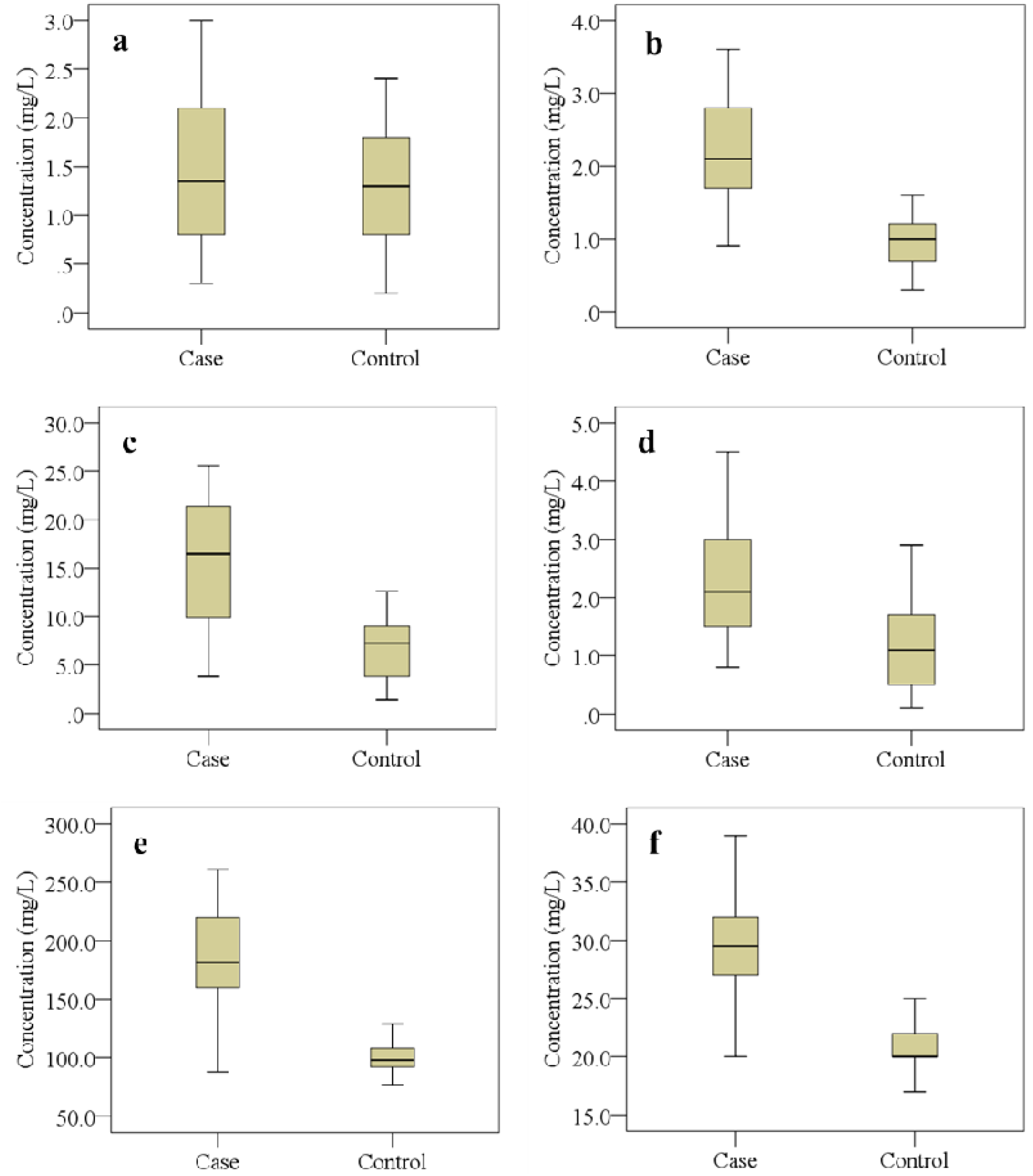
The changes of serum trace elements and macro-minerals (mg/L) levels in the study population. Graphs showing the median, maximum, and minimum value range. **a** Zinc, **b** Copper, **c** Manganese, **d** Iron, **e** Calcium, **f** Magnesium.

**Fig. 2.**
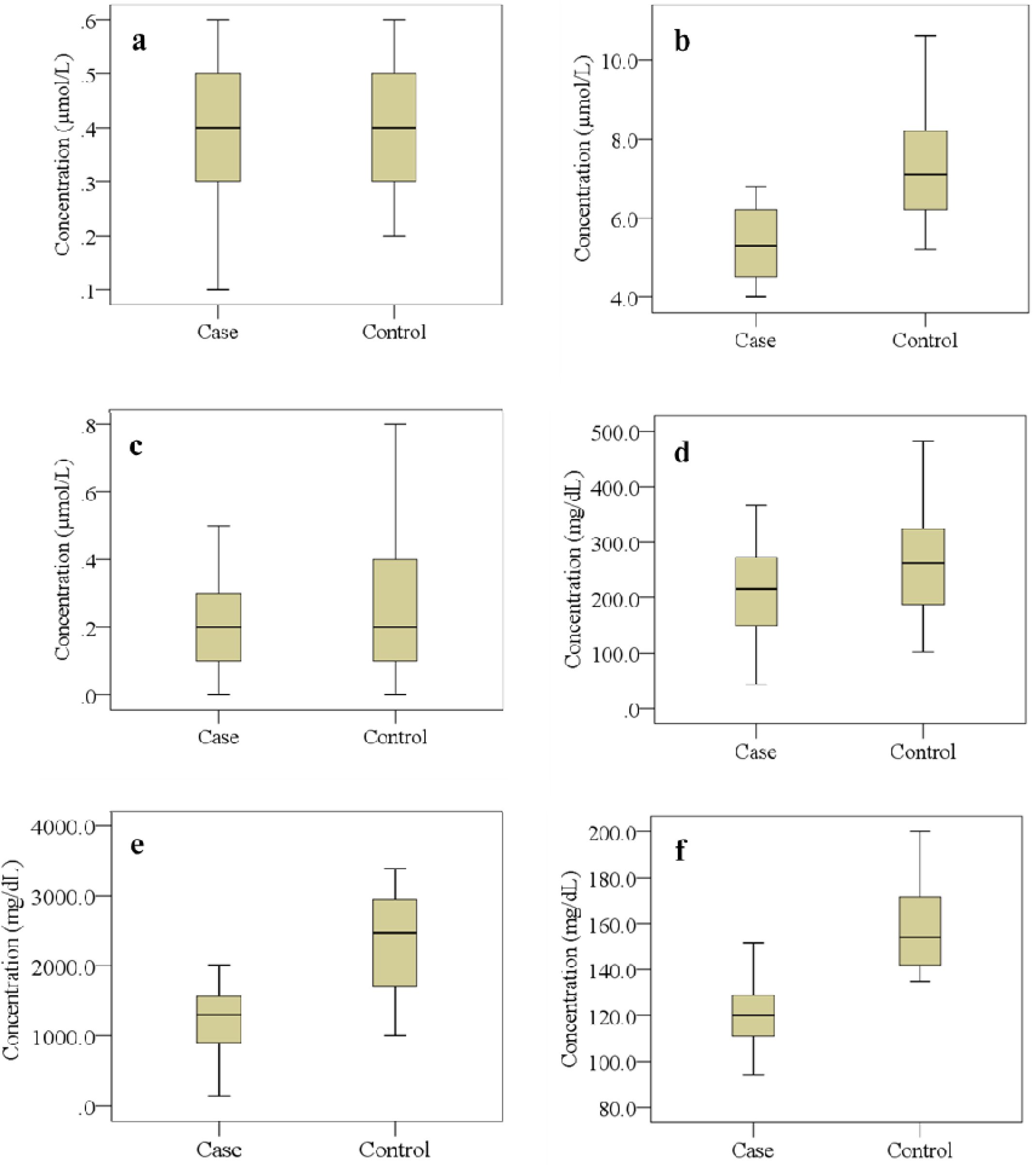
The changes in serum levels of antioxidant vitamins (µmol/L), MDA (µmol/L), and immunoglobulins (mg/dL) levels in the study population. Graphs showing the median, maximum, and minimum value range. **a** Vitamin A, **b** Vitamin E, **c** MDA, **d** Immunoglobulin A, **e** Immunoglobulin G, **f** Immunoglobulin M.

The inter-element relationships of the tested elements between SD patients and HCs were analyzed by Pearson’s correlation test and presented in Table 3. From correlation study, we found a significant positive correlation between Zn and Ca (*r* = 0.365, *p* = 0.009) in SD patients. Also, serum Ca and Mg levels are observed positively correlated with each other (*r* = 0.783, *p* = 0.001) in HCs. However, a significant negative correlation was detected between serum levels of Cu and Ca (*r* = -0.298, *p* = 0.035) in SD patients but no such relation was seen in HCs.

**Table 3.** Study of inter-element correlations in SD patients and healthy controls

| Correlation<br>parameters | SD patients ($n = 75$) | | Healthy controls ($n = 76$) | |
| --- | --- | --- | --- | --- |
| | $r$ | $p$ | $r$ | $p$ |
| Zn and Cu | -0.182 | 0.207 | -0.272 | 0.056 |
| Zn and Mn | 0.001 | 0.996 | 0.003 | 0.887 |
| Zn and Fe | 0.222 | 0.121 | 0.045 | 0.758 |
| Zn and Ca | 0.365 | 0.009** | 0.179 | 0.213 |
| Zn and Mg | 0.231 | 0.107 | -0.127 | 0.379 |
| Cu and Mn | 0.092 | 0.525 | 0.022 | 0.878 |
| Cu and Fe | 0.237 | 0.097 | -0.016 | 0.914 |
| Cu and Ca | -0.298 | 0.035* | -0.095 | 0.512 |
| Cu and Mg | -0.072 | 0.617 | -0.029 | 0.842 |
| Mn and Fe | -0.125 | 0.385 | 0.005 | 0.973 |
| Mn and Ca | -0.127 | 0.380 | 0.116 | 0.422 |
| Mn and Mg | -0.041 | 0.779 | 0.054 | 0.708 |
| Ca and Mg | 0.277 | 0.052 | 0.783 | < 0.001* |

\* $p$ is significant at the 0.05 level; Negative values indicate the opposite correlation.
$r$ correlation coefficient, $p$ significance, SD seborrheic dermatitis.

## Discussion

Analysis of serum trace elements indicated that the serum concentration of Cu, Mn, Fe, Ca, and Mg was increased significantly in SD patients compared to HCs. The role of Zn in SD is obscure since no significant change was found, which is supported by the previous study [21]. Mn is involved in cell differentiation and proliferation; an over-expression of manganese superoxide dismutase enzyme may involve in malignant melanoma [37, 38]. Mn concentrations in serum may catalyze the conversion of H_2_O_2_ to potent hydroxyl radical which could lead to oxidative damage [13] which merely represents an epiphenomenon related to the pathogenesis of SD. Excess amount of Fe was observed in SD patients which might be responsible for inducing free radical-mediated oxidative damage [39]. Ca is also important for the activation of different enzymes, formation of bone, blood clotting system, and neuromuscular excitability [4, 40]. We found the significantly higher levels of Ca in SD patients, whereas one study has confirmed that the elevated serum levels of two calcium-binding proteins are associated with abnormal keratinocyte differentiation in psoriasis [8]. As a MM, Ca forms the action of the second messenger and regulates keratinocyte proliferation and differentiation [41, 42]. The immune response can be altered from several aspects by Cu deficiency or high intakes over longer periods [43, 44]. The present study results along with the previous findings suggest that increased levels of Cu, Mn, Fe, Ca, and Mn may lead to the imbalance in elemental homeostasis which may cause oxidative damage to biomolecules in the skin. Moreover, we have found some significant inter-element relationships in SD patients. According to the present study results, in SD patients, serum Ca levels are positively influenced by the serum levels of Zn whereas serum Ca levels are negatively influenced by the serum Cu levels.

Antioxidant vitamins are considered to be important factors for maintaining the balance between ROS production and elimination [45]. Although ROS is produced essentially in biological processes, it is harmful to the body as it causes cell damage by a free radical chain reaction. Antioxidants inhibit and scavenge free radicals and thereby counteract their effects [7, 46]. The present study showed significantly higher levels of MDA and lower levels of vitamin E, though the changes in vitamin A levels were not significant in SD patients compared to HCs. These results specify cell and tissue damage due to oxidative stress by binding of free radicals with membrane lipids, DNA, and other intracellular components. Our study results are supported by a previous study where significantly lower levels of vitamin E were reported in SD patients compared to HCs [47]. In the scalp, SD patients reported significantly higher levels of MDA when compared to HCs [4]. According to the present findings and previous observations, it implies that elevated MDA and decreased antioxidant vitamin levels may serve as a possible causative factor for the development of SD.

The prevalence of SD is higher (83%) among the population who undergone organ transplantation, who has AIDS, chronic alcoholic pancreatitis, and various malignancies [48]. This observation suggests that the immune system is important for the pathogenesis of SD. The present investigation observed the significantly decreased levels of serum IgA, IgG, and IgM in SD patients compared to HCs. Some previous studies were also reported similar observations [28, 49]. Moreover, SD has been observed more commonly in immunosuppressed patients [48]. From the above discussion, we can say that any alterations in cellular TEs, MMs, antioxidant vitamins, MDA, and immunoglobulins have unfavorable functional consequences on the immune system. Therefore, changes in these parameters in serum levels potentially linked with the etiology of SD.

The present study has some limitations. We did not investigate inflammatory, pro-inflammatory, and anti-inflammatory serum cytokines. ROS can cause different types of oxidative damages and enhance the secretion of these cytokines in SD. Additionally, it would be better if we could measure the levels of analyzed parameters after remission of SD to produce a more valued conclusion. Thus, future investigations in a similar field overcoming the above issues are suggested for a better understanding of the pathophysiology of SD. Despite these drawbacks, it is expected that this work will play an important role to develop new pathological tools for the proper diagnosis and management of SD patients.

## Conclusion

The present study suggests the possible correlations of altered serum TEs, MMs, MDA, antioxidant vitamins, and immunoglobulins with the pathophysiology of SD. These findings may assist in the understanding of the biochemical basis and etiopathological mechanisms of SD. Moreover, macro and micronutrients, immunoglobulins, and multivitamins would assist thee clinical management of SD patients. The alterations of these parameters in SD patients may be the consequence, and not the cause, of disease. However, further detailed and more comprehensive studies designed on a longitudinal basis are needed to discover exactly whether the altered parameters are the cause or consequence of the disease process in SD.

## Data Availability

Data supporting our findings are available from the corresponding author on reasonable request.

## Abbreviations

AAD: American Academy of Dermatology;
BMI: Body mass index;
BMI: Body mass index;
BSMRAU: Bangabandhu Sheikh Mujibur Rahman Agricultural University;
CED: Chronic energy deficiency;
FAAS: Flame atomic absorption spectroscopy;
GFAAS: Graphite furnace atomic absorption spectrometry;
Ig: Immunoglobulin;
MDA: Malondialdehyde;
MM: Macro-minerals;
ROS: Reactive oxygen species;
SD: Seborrheic dermatitis;
SEM: Standard error mean;
SOD: Superoxide dismutase;
SPSS: Statistical package for the social sciences;
TBA: Thiobarbituric acid;
TE: Trace element.

## Acknowledgment

All the authors are highly grateful to all the staff and members of Ad-din Medical College Hospital, Dhaka. The authors are also thankful for the laboratory support of the Department of Soil Science, Faculty of Agriculture, Bangabandhu Sheikh Mujibur Rahman Agricultural University (BSMRAU), Salna, Gazipur, Bangladesh.

## Authors’ contributions

IJ and MRI Conceived, designed, conducted, and analyzed the experiment, performed computational analyses, and data editing, wrote, and revised the manuscript. MRI and ZN Analyzed the experiment, wrote, and revised the manuscript. RA and SMMR Diagnosed and evaluate the study subjects. AH and MSI Supervised the whole work and gave important intellectual content in the manuscript. All authors have read and approved the final version of this manuscript

## Availability of data and materials

Data supporting our findings are available from the corresponding author on reasonable request.

## Ethics approval and consent to participate

The study protocol was approved by the ethical review committee of the department of dermatology and venereology, Ad-din Medical College Hospital, Dhaka, Bangladesh. All the participants were well informed about the purpose of the study and written consent was obtained from each of them to participate.

## Consent for publication

Not applicable. ’

